# Influenza vaccine effectiveness against medically attended outpatient illness, United States, 2023–24 season

**DOI:** 10.1101/2024.10.29.24316377

**Authors:** Jessie R. Chung, Ashley M. Price, Richard K. Zimmerman, Krissy Moehling Geffel, Stacey L. House, Tara Curley, Karen J. Wernli, C. Hallie Phillips, Emily T. Martin, Ivana A. Vaughn, Vel Murugan, Matthew Scotch, Elie A. Saade, Kiran A. Faryar, Manjusha Gaglani, Jason D. Ramm, Olivia L. Williams, Emmanuel B. Walter, Marie Kirby, Lisa M. Keong, Rebecca Kondor, Sascha R. Ellington, Brendan Flannery, US Flu VE Network Investigators

## Abstract

**Background:** The 2023–24 U.S. influenza season was characterized by a predominance of A(H1N1)pdm09 virus circulation with co-circulation of A(H3N2) and B/Victoria viruses. We estimated vaccine effectiveness (VE) in the United States against mild-to-moderate medically attended influenza illness in the 2023–24 season.

**Methods:** We enrolled outpatients aged ≥8 months with acute respiratory illness in 7 states. Respiratory specimens were tested for influenza type/subtype by reverse-transcriptase polymerase chain reaction (RT-PCR). Influenza VE was estimated with a test-negative design comparing odds of testing positive for influenza among vaccinated versus unvaccinated participants. We estimated VE by virus sub-type/lineage and A(H1N1)pdm09 genetic subclades.

**Results:** Among 6,589 enrolled patients, 1,770 (27%) tested positive for influenza including 796 A(H1N1)pdm09, 563 B/Victoria, and 323 A(H3N2). Vaccine effectiveness against any influenza illness was 41% (95% Confidence Interval [CI]: 32 to 49): 28% (95% CI: 13 to 40) against influenza A(H1N1)pdm09, 68% (95% CI: 59 to 76) against B/Victoria, and 30% (95% CI: 9 to 47) against A(H3N2). Statistically significant protection against any influenza was found for all age groups except adults aged 50–64 years. Lack of protection in this age group was specific to influenza A-associated illness. We observed differences in VE by birth cohort and A(H1N1)pdm09 virus genetic subclade.

**Conclusions:** Vaccination reduced outpatient medically attended influenza overall by 41% and provided protection overall against circulating influenza A and B viruses. Serologic studies would help inform differences observed by age groups.

**Key Points:** Influenza vaccine reduced the risk of outpatient illness due to influenza during the 2023–24 season. Protection varied by age group and influenza virus type.

## Introduction

Influenza vaccination is a proven safe and effective strategy for mitigating the burden of influenza-associated morbidity. In the United States, influenza vaccination continues to be recommended annually for all persons aged ≥6 months who do not have contraindications [1, 2]. US adults aged ≥65 years are preferentially recommended to receive a higher dose or adjuvanted vaccine. During the U.S. 2022–23 influenza season, vaccination was estimated to have prevented 5.9 million illnesses, 2.9 million medical visits, 64,000 hospitalizations, and 3,600 deaths associated with influenza [3]. Despite the known benefits of influenza vaccination, coverage rates are far below Healthy People 2030 target of achieving 70% coverage for all age groups [4, 5]. In the 2023–24 influenza season, an estimated 49% of adults and 54% of children were vaccinated against influenza [6, 7].

The 2023–24 influenza season in the United States was characterized by a predominance of A(H1N1)pdm09 virus circulation with co-circulation of A(H3N2) and B/Victoria viruses [8]. Three-quarters of sequenced A(H1N1)pdm09 viruses belonged to hemagglutinin (HA) clade 6B.1A.5a subclade 2a.1 (which includes the 2023–24 vaccine reference strain), with the remaining quarter belonging to clade 6B.1A.5a subclade 2a. Co-circulation of two genetic subclades of the predominant A(H1N1)pdm09 virus provided the opportunity to investigate clade-specific protection. Mid-season data through January 2023 from four U.S. vaccine effectiveness platforms showed significant protection against outpatient and inpatient influenza illness [9]. Here we report updated vaccine effectiveness (VE) for the 2023–24 season against mild-to-moderate outpatient medically attended influenza overall by influenza A subtype, influenza B lineage, and age group.

## Methods

### Study population

The US Flu VE Network has been described previously [10, 11]. In brief, the public health surveillance network enrolls participants aged 8 months or older with an acute respiratory illness (ARI) ≤7 days duration including new or worsening cough at sites in Arizona, Michigan, Missouri, Ohio, Pennsylvania, Texas, and Washington. Duke University, North Carolina, served as the Network’s data coordinating institution. Enrollment began at each site based on local evidence of increasing influenza activity. Trained staff interviewed participants or their parent/guardian to obtain demographic and clinical information and inquired about receipt of current season influenza vaccination. Underlying chronic health conditions were defined as electronic medical record documentation within the previous year of medical encounters with International Classification of Diseases (ICD-10) codes for high-risk conditions according to the Advisory Committee on Immunization Practices (ACIP) [1]. Enrollment dates ranged from October 1, 2023–April 30, 2024 (**Supplemental Table 1**). The study protocol was reviewed by US Centers for Disease Control and Prevention (CDC), determined to be public health surveillance, and conducted consistent with applicable federal law and CDC policy (45 CFR 46.102(l)(2)).

### Influenza vaccination

Northern Hemisphere 2023–24 quadrivalent influenza vaccines included two influenza A antigens and two influenza B antigens. Egg-based vaccines were recommended to include A/Victoria/4897/2022-like (H1N1pdm09-like genetic subclade 2a.1) and cell-culture- (ccIIV4) or recombinant-based (RIV) vaccines were recommended to include A/Wisconsin.67/2022-like (H1N1pdm09-like genetic subclade 2a.1). Each of the three vaccine types were recommended to also include A/Darwin/9/2021-like (A/H3N2-like genetic clade 3C.2a1b.2a.2), B/Austria/1359417/2021-like (B/Victoria-like genetic clade V1A.3a.2), and B/Phuket/3073/2013- like (B/Yamagata genetic clade Y3) [12]. Participant influenza vaccination status was determined combining data from electronic health records (EHR), state immunization information systems, and plausible self-report. A plausible self-reported influenza vaccination was a dose reported by the participant (or parent/guardian of participant) that was received more than 14 days prior to illness onset and included a vaccination provider location. All participants were classified as vaccinated if they had received one or more doses of any licensed influenza vaccine product after July 1, 2023.

### Laboratory methods

Participants had nasal and oropharyngeal swabs obtained (nasal only for children younger than 2 years of age). Specimens were tested for influenza (including type and influenza A subtype) and SARS-CoV-2 viruses by molecular assays at site laboratories. Participants who tested positive for influenza were designated as cases and participants who tested negative for influenza and SARS-CoV-2 were designated non-cases (controls).

Influenza-positive specimens were genetically characterized using next generation sequencing locally (one site, AZ) or at CDC (six sites). Each sample’s HA clade was based on its consensus sequence [13].

### Statistical analysis

We excluded the following from the primary analyses: participants with inconclusive influenza test results, participants who tested positive for SARS-CoV-2 [14], and participants who had been vaccinated less than 14 days prior to illness onset. Influenza vaccine effectiveness was estimated with a test-negative design comparing odds of testing positive for influenza among vaccinated versus unvaccinated participants [15]. VE is expressed as (1 – OR) x 100, where OR is the adjusted odds ratio for influenza among vaccinated persons vs unvaccinated persons from logistic regression models. Models were adjusted a priori for study site, participant age (group for all-ages estimates and year of age for age group-specific estimates) at enrollment, presence of ≥1 underlying health condition, and month of illness onset.

Additional covariates (i.e., sex, race and ethnicity, self-rated general health status, and interval between onset and enrollment) were considered using a threshold for inclusion of ≥5% change in the OR. Estimates include 95% confidence intervals where exclusion of 0% indicates statistically significant VE.

We estimated VE by influenza type/subtype and by age group (as 8 months – 8 years, 9–17 years, 18–49 years, 50–64 years, and ≥65 years). Age group-specific estimates were combined when models did not converge. We also examined VE against A(H1N1)pdm09 among people born 1958–1979 (ages 45–65 years) because this cohort has been associated with lower effectiveness against A(H1N1)pdm09 in prior seasons [16]. We estimated age group-specific VE against any influenza by time since vaccination using intervals of 14–60 days, 60– 120 days, and >120 days since vaccination compared with unvaccinated participants.

## Results

### Participant characteristics

From October 1, 2023, through April 30, 2024, we enrolled 9,061 participants who presented to outpatient and urgent care clinics and emergency departments for acute respiratory illness. We excluded 2,472 participants from primary analyses; over half of excluded participants (1,294, 52%) tested positive for SARS-CoV-2 (**Supplemental Figure 1**). Of the included 6,589 participants, 1,770 (27%) tested positive for influenza (Table 1): 796 (45%) for A(H1N1)pdm09, 563 (32%) for influenza B/Victoria, 323 (18%) for A(H3N2), and 112 (6%) for influenza A viruses of undetermined subtype; 20 (1%) participants had influenza A and B virus coinfections, and 4 (<1%) had influenza A(H1N1)pdm09 and A(H3N2) coinfections. Notably, a smaller proportion of B/Victoria cases occurred in participants aged ≥50 years compared to influenza A subtype viruses (**Supplemental Figure 2**). The number of influenza-positive participants enrolled per week peaked in late January 2024 (**Supplemental Figure 3**). A total of 2,489 (38%) participants were considered vaccinated for influenza for the 2023–24 season. Of these, 403 (16%) had no documented immunization record of 2023–24 influenza vaccine but had plausible self-reported receipt.

**Table 1.** Characteristics of participants enrolled in the US Influenza Vaccine Effectiveness Network for the 2023–24 influenza season ^a^.

|  | RT-PCR Test result status |  |  |  |  |  | Influenza Vaccination status |  |  |
| --- | --- | --- | --- | --- | --- | --- | --- | --- | --- |
|  | Total | Influenza-positive |  | Influenza-negative |  |  | Vaccinated |  |  |
| Characteristic | No. | No. | Column % | No. | Column % | p-value | No. | Row % | p-value |
| *Overall* | 6589 | 1770 | 100 | 4819 | 100 |  | 2489 | 37 |  |
| *Study site* |  |  |  |  |  | <0.01 |  |  | <0.01 |
| Arizona | 577 | 206 | 12 | 371 | 8 |  | 74 | 13 |  |
| Michigan | 367 | 82 | 5 | 285 | 6 |  | 183 | 50 |  |
| Missouri | 779 | 345 | 19 | 434 | 9 |  | 225 | 29 |  |
| Ohio | 766 | 257 | 15 | 509 | 11 |  | 229 | 30 |  |
| Pennsylvania | 1178 | 353 | 20 | 825 | 17 |  | 512 | 43 |  |
| Texas | 1798 | 351 | 20 | 1447 | 30 |  | 739 | 41 |  |
| Washington | 1124 | 176 | 10 | 948 | 20 |  | 527 | 47 |  |
| *Sex* <sup>b</sup> |  |  |  |  |  | 0.03 |  |  | <0.01 |
| Female | 3972 | 1030 | 58 | 2942 | 61 |  | 1570 | 40 |  |
| Male | 2601 | 738 | 42 | 1863 | 39 |  | 915 | 35 |  |
| *Age group (years)* |  |  |  |  |  | <0.01 |  |  | <0.01 |
| 8 months–8 years | 1327 | 397 | 22 | 930 | 19 |  | 342 | 26 |  |
| 9–17 | 613 | 229 | 13 | 384 | 8 |  | 147 | 24 |  |
| 18–49 | 2699 | 774 | 44 | 1925 | 40 |  | 860 | 32 |  |
| 50–64 | 1013 | 228 | 13 | 785 | 16 |  | 486 | 48 |  |
| ≥65 | 937 | 142 | 8 | 795 | 16 |  | 654 | 70 |  |
| *Race and ethnicity* <sup>c</sup> |  |  |  |  |  |  |  |  |  |
| American Indian or Alaska Native | 119 | 22 | 1 | 97 | 2 | 0.04 | 32 | 27 | 0.01 |
| Asian | 344 | 107 | 6 | 237 | 5 | 0.07 | 141 | 41 | 0.21 |
| Black or African American | 1861 | 601 | 34 | 1260 | 26 | <0.01 | 479 | 26 | <0.01 |
| Native Hawaiian or Other Pacific Islander | 69 | 21 | 1 | 48 | 1 | 0.50 | 15 | 22 | 0.01 |
| White | 4229 | 1014 | 57 | 3215 | 67 | <0.01 | 1823 | 43 | <0.01 |
| Hispanic or Latino | 1015 | 283 | 16 | 732 | 15 | 0.44 | 290 | 29 | <0.01 |
| *Self-rated general health status* <sup>d</sup> |  |  |  |  |  | <0.01 |  |  | <0.01 |
| Excellent | 1656 | 529 | 30 | 1127 | 23 |  | 508 | 31 |  |
| Very good | 2258 | 616 | 35 | 1642 | 34 |  | 855 | 38 |  |
| Good | 1849 | 461 | 26 | 1388 | 29 |  | 786 | 43 |  |
| Fair/Poor | 809 | 161 | 9 | 648 | 13 |  | 337 | 42 |  |
| *Underlying chronic health conditions* <sup>e</sup> |  |  |  |  |  | <0.01 |  |  | <0.01 |
| None | 3820 | 1222 | 69 | 2598 | 54 |  | 1086 | 28 |  |
| ≥1 | 2769 | 548 | 31 | 2221 | 46 |  | 1403 | 51 |  |
| *Illness onset to enrollment (days)* |  |  |  |  |  | <0.01 |  |  | <0.01 |
| <3 | 2233 | 723 | 41 | 1510 | 31 |  | 767 | 34 |  |
| 3–4 | 2030 | 606 | 34 | 1424 | 30 |  | 704 | 35 |  |
| 5–7 | 1201 | 264 | 15 | 937 | 19 |  | 493 | 41 |  |

<sup>a</sup> Included patients with medically attended acute respiratory illness enrolled in the US Influenza Vaccine Effectiveness network between October 2023 through April 2024.
<sup>b</sup> Sex was missing for 16 participants.
<sup>c</sup> Participants self-identified all options that applied and could have chosen more than one. Race and ethnicity were missing for 69 participants.
<sup>d</sup> Self-rated general health status was missing for 17 participants.
<sup>e</sup> Underlying chronic health conditions included chronic cardiac diseases and circulatory diseases, chronic pulmonary diseases, diabetes mellitus, chronic renal disease, hemoglobinopathies, immunosuppressive disorders, malignancy, metabolic diseases, liver diseases, neurological/musculoskeletal conditions, cerebrovascular disease, morbid obesity, and endocrine disorders.

Influenza-positive cases were more likely to be male, of younger age, identify themselves as Black or African American, report better general health, and seek care earlier compared to test-negative controls; cases were less likely to have an underlying health condition than controls (**Table 1**). The proportion of participants vaccinated differed by study site, sex, age, race and ethnicity, general health status, presence of an underlying health condition, and number of days between illness onset to enrollment (**Table 1**). A total of 1,513 (61%) vaccinated participants had a documented vaccine type. Of those with known vaccine type, 1,362 (90%) received egg-based vaccines and 151 (10%) received non-egg-based vaccines (123 ccIIV4, 28 RIV4). Among 654 vaccinated adults aged ≥65 years, 492 (75%) had a known vaccine type. Of those, 467 (95%) received a preferentially recommended vaccine: 281 (57%) received high-dose inactivated influenza vaccine and 186 (38%) received adjuvanted influenza vaccine; none received RIV4.

### Genetic characterization

A total of 1,185 viruses were successfully sequenced: 612 A(H1N1)pdm09, 340 B/Victoria, and 233 A(H3N2). A(H1N1)pdm09 viruses belonged to two genetic subclades: 474 (77%) to 6B.1A.5a.2a.1 (2a.1), the same genetic subclade as the 2023–24 A(H1N1)pdm09 vaccine reference virus, and 138 (23%) to 6B.1A.5a.2a (2a). The 2a.1 genetic subclade predominated among sequenced A(H1N1)pdm09 viruses throughout the enrollment period (**Supplemental Figure 4**). All B/Victoria viruses belonged to genetic clade V1A.3a.2, and all A(H3N2) viruses belonged to genetic subclade 2a.3a.1.

### Vaccine effectiveness

From October 2023 through April 2024, the overall adjusted VE against medically attended outpatient influenza A and B viruses was 41% (95% confidence interval [CI]: 32 to 49) (Table 2). Estimated VE was 28% (95%CI: 13 to 40) against influenza A(H1N1)pdm09, 68% (95%CI: 59 to 76) against influenza B/Victoria, and 30% (95%CI: 9 to 47) against A(H3N2). VE against A(H1N1)pdm09 varied by age group; VE was lowest, offering no protection against medically attended outpatient illness, among adults aged 50–64 years (-8%, 95%CI: -62 to 28) and was highest among children aged 8 months – 8 years (59%, 95%CI: 31 to 77). Among adults born in a specific birth cohort (birth years 1958–1979), we observed no protection in terms of medically attended illness against A(H1N1)pdm09-associated influenza (2% VE (95%CI: -38 to 30)). Among participants of all ages, VE was higher against influenza A(H1N1)pdm09 genetic subclade 2a viruses (59%, 95%CI: 35 to 75) compared to genetic subclade 2a.1 viruses (23%, 95%CI: 4 to 39) (Figure 1). When stratified by age group, the greatest difference in VE between the two genetic subclade was observed among participants aged 18–64 years. VE against A(H1N1)pdm09 genetic subclade 2a.1 viruses in this age group was 13% (95%CI: -16 to 35) versus 61% (95%CI: 30 to 80) against viruses in the 2a genetic subclade.

**Figure 1.**
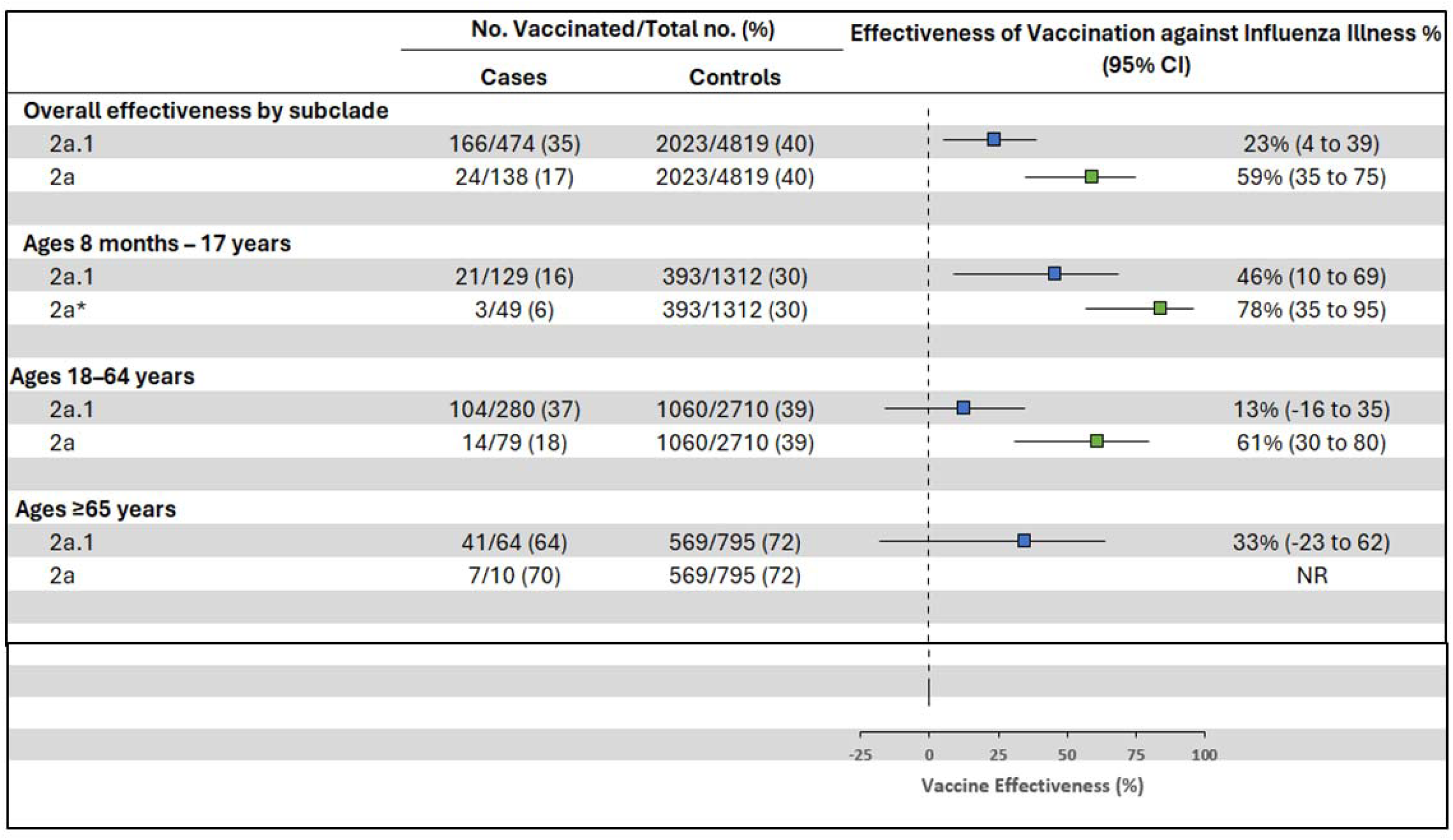
Adjusted^a^ vaccine effectiveness against outpatient influenza A(H1N1)pdm09 genetic subclade-associated illness visits among patients aged ≥8 months enrolled at US Influenza Vaccine Effectiveness Network sites, October 2023 through April 2024. CI, confidence interval; NR, not reported due to small sample size ^a^ Models adjusted for study site, age, presence of ≥1 underlying health condition, and month of illness onset. 95% confidence intervals that exclude 0% are considered statistically significant *Unadjusted due to small sample size

**Table 2.**
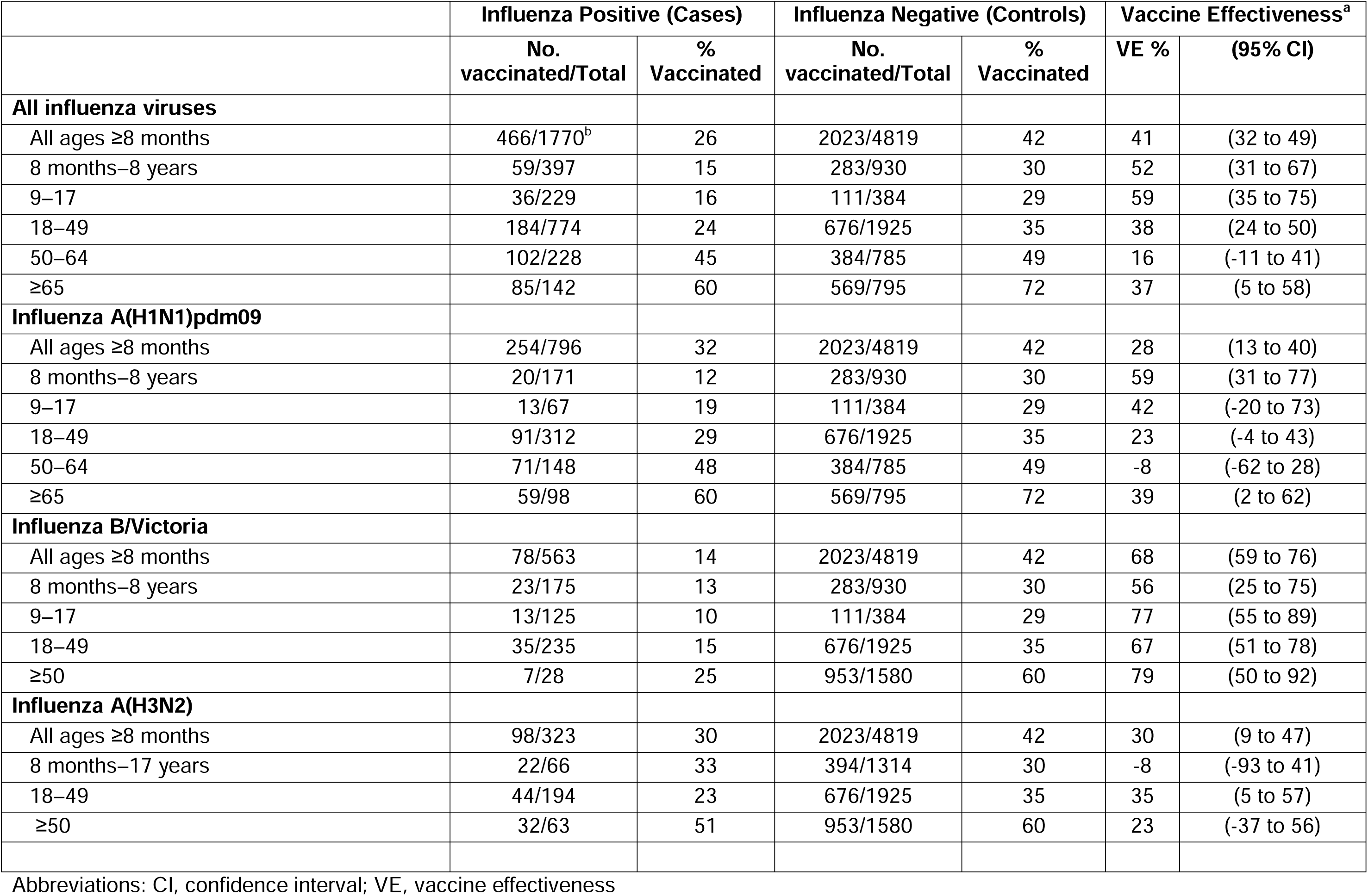
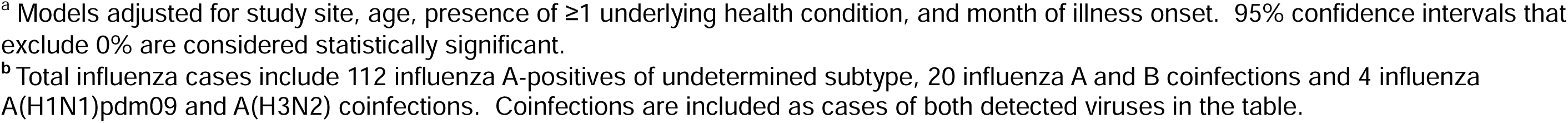
Adjusted vaccine effectiveness against outpatient influenza-associated illness visits among patients aged ≥8 months enrolled at US Influenza Vaccine Effectiveness Network sites, October 2023 through April 2024.

|  | Influenza Positive (Cases) |  | Influenza Negative (Controls) |  | Vaccine Effectiveness<sup>a</sup> |  |
| --- | --- | --- | --- | --- | --- | --- |
|  | No. vaccinated/Total | % Vaccinated | No. vaccinated/Total | % Vaccinated | VE % | (95% CI) |
| **All influenza viruses** |  |  |  |  |  |  |
| All ages ≥8 months | 466/1770<sup>b</sup> | 26 | 2023/4819 | 42 | 41 | (32 to 49) |
| 8 months–8 years | 59/397 | 15 | 283/930 | 30 | 52 | (31 to 67) |
| 9–17 | 36/229 | 16 | 111/384 | 29 | 59 | (35 to 75) |
| 18–49 | 184/774 | 24 | 676/1925 | 35 | 38 | (24 to 50) |
| 50–64 | 102/228 | 45 | 384/785 | 49 | 16 | (-11 to 41) |
| ≥65 | 85/142 | 60 | 569/795 | 72 | 37 | (5 to 58) |
| **Influenza A(H1N1)pdm09** |  |  |  |  |  |  |
| All ages ≥8 months | 254/796 | 32 | 2023/4819 | 42 | 28 | (13 to 40) |
| 8 months–8 years | 20/171 | 12 | 283/930 | 30 | 59 | (31 to 77) |
| 9–17 | 13/67 | 19 | 111/384 | 29 | 42 | (-20 to 73) |
| 18–49 | 91/312 | 29 | 676/1925 | 35 | 23 | (-4 to 43) |
| 50–64 | 71/148 | 48 | 384/785 | 49 | -8 | (-62 to 28) |
| ≥65 | 59/98 | 60 | 569/795 | 72 | 39 | (2 to 62) |
| **Influenza B/Victoria** |  |  |  |  |  |  |
| All ages ≥8 months | 78/563 | 14 | 2023/4819 | 42 | 68 | (59 to 76) |
| 8 months–8 years | 23/175 | 13 | 283/930 | 30 | 56 | (25 to 75) |
| 9–17 | 13/125 | 10 | 111/384 | 29 | 77 | (55 to 89) |
| 18–49 | 35/235 | 15 | 676/1925 | 35 | 67 | (51 to 78) |
| ≥50 | 7/28 | 25 | 953/1580 | 60 | 79 | (50 to 92) |
| **Influenza A(H3N2)** |  |  |  |  |  |  |
| All ages ≥8 months | 98/323 | 30 | 2023/4819 | 42 | 30 | (9 to 47) |
| 8 months–17 years | 22/66 | 33 | 394/1314 | 30 | -8 | (-93 to 41) |
| 18–49 | 44/194 | 23 | 676/1925 | 35 | 35 | (5 to 57) |
| ≥50 | 32/63 | 51 | 953/1580 | 60 | 23 | (-37 to 56) |

Abbreviations: CI, confidence interval; VE, vaccine effectiveness
<sup>a</sup> Models adjusted for study site, age, presence of $\geq 1$ underlying health condition, and month of illness onset. 95% confidence intervals that exclude 0% are considered statistically significant.
<sup>b</sup> Total influenza cases include 112 influenza A-positives of undetermined subtype, 20 influenza A and B coinfections and 4 influenza A(H1N1)pdm09 and A(H3N2) coinfections. Coinfections are included as cases of both detected viruses in the table.

The median time between vaccination and illness onset was 94.5 days [interquartile range (IQR) 58–134 days]. The interval was shorter among children aged 8 months – 17 years (median 83 days [IQR: 51, 128]) than among adults aged 18–64 years (median 95 days [IQR: 56, 134]) and adults aged ≥65 years (median 99.5 days [IQR: 64.5, 142]). Half of vaccinated adults aged ≥65 years were vaccinated by October 4, 2023, compared to October 12, 2023 for adults aged 18–49 years and October 21, 2023 for children and adolescents aged 8 months – 17 years. Overall and for each age group, VE point estimates were highest among people vaccinated 14–59 days prior to illness onset compared to those with longer intervals although confidence intervals largely overlapped (**Supplemental Table 2**). We observed protection against outpatient influenza through 120 or more days after vaccination among both children and adults aged <64 years. Among adults aged ≥65 years, no protection was observed ≥60 days after vaccination.

### Sensitivity analyses

VE calculated using different sources of influenza vaccination data were within 10 percentage points of overall adjusted VE against any influenza combining data sources. VE against any influenza using self- or parent/guardian-reported influenza vaccination was 32% (95%CI: 21 to 41); VE using EHR-documented data only was 44% (95%CI: 35 to 52) (**Supplemental Table 3**). Sensitivity analyses varying inclusion criteria for analysis were all within 4 percentage points of the overall adjusted VE against any influenza reported in the primary analysis.

## Discussion

In a season with co-circulation of A(H1N1)pdm09, A(H3N2), and B/Victoria influenza viruses, we estimated that influenza vaccination reduced the risk of outpatient medically attended influenza by 41%. We found significant protection overall against A(H1N1)pdm09, B/Victoria, and A(H3N2) viruses. Observed vaccine protection varied against the two A(H1N1)pdm09 genetic subclades that circulated in the United States; lower protection was observed against the predominant A(H1N1)pdm09 virus (genetic subclade 2a.1) that was included in the 2023–24 vaccine. Protection against any influenza was greatest 14–59 days after vaccination. Protection was observed more than 120 days after vaccination for participants aged <65 years.

We report differences in effectiveness by age group and influenza A virus subtype, that may contribute to our understanding of antigenic imprinting with A(H1N1) viruses. Specifically, we observed no significant protection against medically attended illness among adults aged 18– 64 years against influenza A(H1N1)pdm09-related illness. Reduced protection in this age category appeared limited to A(H1N1)pdm09 viruses in the 2a.1 genetic subclade. Restricting further by birth cohort, we observed no protection among participants born 1958–1979, the same birth cohort previously associated with reduced VE against A(H1N1)pdm09 viruses [17–19]. Individuals in this birth cohort were likely exposed to influenza A(H2N2) or A(H3N2) before A(H1N1) and were first exposed to A(H1N1) viruses after this subtype re-emerged in 1977 [20]. This birth cohort has experienced lower VE against A(H1N1)pdm09 clade 6B.1 viruses compared to other cohorts [17, 18]. Serologic studies of vaccine response would help to understand potential birth cohort effects and inform development of improved influenza vaccines that might overcome these effects. The A(H1N1)pdm09 vaccine antigen will continue to be an A/Wisconsin/67/2022-like virus antigen from the 2a.1 genetic subclade for the 2024–25 Northern Hemisphere season [21].

Results from this study of VE against medically attended ARI in outpatient or ambulatory settings are consistent with findings from active enrollment and electronic health record (EHR)- based surveillance platforms in different populations and settings in the 2023–24 season. While the US Flu VE Network estimates VE against medically attended outpatient illness, some evidence suggests that influenza vaccination could attenuate influenza disease severity [22].

Mid-season estimates of VE from four US VE studies conducted in outpatient and inpatient settings demonstrated early protection against illness and hospitalizations; overall VE against any influenza ranged from 52–67% among children and adolescents and 33–49% among adults [9]. Estimates were similar against both mild-to-moderate outpatient illness and influenza-associated hospitalizations. A previous analysis comparing 25 paired VE estimates in outpatient and inpatient settings found similar levels of protection against mild-to-moderate and more severe influenza [23]. Further, estimates from the US-based VISION EHR-based surveillance platform [24] found similar VE against influenza A and B in outpatient (emergency department and urgent care clinics) and inpatient settings consistent with US Flu VE Network estimates among enrolled outpatients. Among VE studies conducted outside the United States, mid-season estimates from Canada reported higher overall effectiveness against A(H1N1)pdm09 (63%, 95% CI: 51 to 72) in a setting of approximately equal detections of genetic subclades 2a.1 and 2a [25]. VE was lower against 2a.1 viruses (56%, 95%CI: 33 to 71) than against 2a viruses (67%, 95% CI: 48 to 80); though CIs overlapped. Similar trends were reported from European outpatient and inpatient studies [26] and an health administration data-based study in Alberta, Canada [27].

Like other observational studies, this study is subject to several limitations. First, some misclassifications of exposure may have occurred. In a small proportion (16%) of vaccinated participants, we relied on self-reported influenza vaccine receipt that was not documented in immunization records. It has been reported that the number of influenza vaccines administered in medical offices has declined by about 32% since the COVID-19 pandemic as people increasingly seek vaccinations from pharmacies [4]. This shift may affect availability of vaccination data in EHRs. VE estimates in sensitivity analyses varying source of vaccination status data were comparable, however. Second, analyses of VE by virus genetic clade/subclade, birth cohort, and time since vaccination were limited by the number of patients enrolled during the influenza season. Finally, while the test negative design reduces potential bias due to differences in healthcare seeking, unmeasured confounding related to study enrollment may have affected VE estimates. However, our findings were robust to changes in inclusion criteria and definitions of exposure status.

The results presented in this study demonstrate an overall benefit of receiving the 2023– 24 influenza vaccine in the United States. As influenza vaccine components are updated, annual studies are needed to assess protections against circulating strains and emergent genetic clades. VE studies may contribute to future vaccine strain selection and improvements in influenza vaccines.

## Data Availability

All data produced in the present study may be made available upon reasonable request to the authors.

## Funding and Conflicts of interest

The US Flu VE Network is funded through a US Centers for Disease Control and Prevention Cooperative Agreement (1U01 IP001180-01, 1U01 IP001181-01, 1U01 IP001182-01, 1U01 IP001184-01, 1U01 IP001189-01, 1U01 IP001191-01, 1U01 IP001193-01, and 1U01 IP001194-01). The University of Pittsburgh site was also supported by National Institutes of Health grant UL1TR001857.

EAS has received grants from Protein Sciences Corporation and consulting fees from Johnson and Johnson. EBW has received research funding from Pfizer, Moderna, Seqirus, Najit Technologies, and Clinetic for the conduct of clinical research studies. He has also received support as an advisor to Vaxcyte and Pfizer consultant to ILiAD Biotechnologies, and DSMB member for Shionogi. ETM has received grants from Merck. RKZ has received grants from Sanofi Pasteur. SLH has received grants from Seegene Inc., Abbott, Healgen, Roche, CorDx, Hologic, Cepheid, Janssen, and Wondfo Biotech. All other authors report nothing to disclose.

## Disclaimer

The findings and conclusions in this report are those of the authors and do not necessarily represent the official position of the Centers for Disease Control and Prevention.

## US Flu VE Network Investigators

Joanna Kramer and Mehal Patel (Phoenix Children’s Hospital); Lora Nordstrom and Mary Mulrow (Valleywise Health); Steven Holland and Temitope O.C. Faleye (Arizona State University); Arnold S. Monto, Donny Hearn III, and Caroline K. Cheng (University of Michigan); Lois Lamerato, Jean Ashley Lava, and Muniza Hossain (Henry Ford Health); Michael Lehmkuhl, Elizabeth Capasso-Gulve, Jamie Mills, Joshua Jackson (Washington University School of Medicine in St. Louis); Natalie A. B. Bontrager, Darren J. Morrow, Wes Rountree, Christopher A. Todd, and Cameron R. Wolfe (Duke University); Robert A. Salata, Kirk A. Stiffler, Jason J. Glagola, Zainab Albar, Christopher Ladikos (University Hospitals of Cleveland); Curtis J. Donskey (Louis Stokes Cleveland VA Medical Center; Mary Patricia Nowalk, GK Balasubramani, Tracey Conti, David A. Figucia, John V. Williams, and Alexandra Weissman (University of Pittsburgh); Spencer Rose, Michael Smith, Leah Odame-Bamfo, Kempapura Murthy, Chandni Raiyani, and Mufaddal Mamawala (Baylor Scott & White Health); Erika Kiniry, Brianna Wickersham, Rachael Doud, and Matthew Nguyen (Kaiser Permanente Washington); Juliana DaSilva, Sydney R. Sheffield, Julia C. Frederick, Malania M. Wilson, Ewelina Lyszkowicz, and Philip Shirk (US Centers for Disease Control and Prevention)

## Acknowledgements

We thank the participants of the US Flu VE Network. The Arizona State University team acknowledges the contributions of Kelly Conard, Bradley Bobbett, Ching-Wen Hou, Lucy Sublasky-Rodriguez, LaRinda Holland, Izamar Garcia, Giavanna Caruth, Alexis Thomas, Gabrielle Hernandez Barrera, Jesus Estrada, Karen M. Yeager, Raquel Salgado, Martine Desulme and Bryan Remuto; The Baylor Scott & White Health team acknowledges the contributions of Amanda McKillop, Britan Fairall, Ashley Graves, Martha Zayed, Marcus Volz, Kimberley Walker, Tora Adams, Jenica Aldas, Vanessa Crumpton, Erica Hayes, Paula Harkins, Victoria Harkins-Walston, Maria Lopez, Arnesa Pepino, Seretha Wallace, Sharla Russell, Briget Da Graca, Muralidhar Jatla, Madhava Beeram, Tresa McNeal, Keith Stone, Arundhati Rao, Manohar Mutnal, Tammy Fisher, Jason Ettlinger, Courtney Shaver, Monica Bennett, Elisa Priest, Jennifer Thomas, Javed Butler and Alejandro Arroliga. Kaiser Permanente Washington further acknowledges the contributions of Kate Castillo, Katherine Chen, Joe Choe, Todd Corder, Jana ffitch, Victor Garcia, Miriam Marcus-Smith, Kathryn Moser, Sherry Kubitz, Stephanie Pimienta, Magali Sanchez, Paul Thottingal, and Brian Williamson. The University of Michigan group acknowledges the contributions of Ishraaq Atkins, Sarita Baranwal, Isabelle Birt, Shane Bole, Jaleesa Clark, Marwa Hussein, Jacoby Jennings, Emiliegh Johnson, Anne Kaniclides, Armanda Kimberly, Regina Lehmann, Janay Patterson, Dolapo Raji, Maanasa Ravishankar, Leo Sullivan, Rachel Truscon. The University of Pittsburgh group acknowledges the contributions of Michael Susick, Theresa M. Sax, Louise H. Taylor, Karen Clarke, Klancie Dauer, Helen D’Agostino, Robert Hickey, Joe Suyama, Myla Stiegler, and Monika Johnson. The Washington University School of Medicine team acknowledges the contributions of the staff of the Emergency Care Research Core including Ian Schamburg, Jessica Zvanut, Minh Nguyen, Jeremy Russell, Nichole Smith, Allison Hardin, Sarah Pruett-Welsh, Vidhya Venkataraman, Taylor Kristicevich, Leah Heath, Elise Blanke, Jamie Garner, Wykita Willis-Lockhart, Heena Khan, and Rianna Robinson.

## Notes

### Author Declarations

The study protocol was reviewed by US Centers for Disease Control and Prevention (CDC), determined to be public health surveillance, and conducted consistent with applicable federal law and CDC policy (45 CFR 46.102(l)(2)).

