## Supplemental Tables and Figures for "Influenza vaccine effectiveness against medically attended outpatient illness, United States, 2023–24 season"

**Supplemental Table 1.** Enrollment dates included by site

| **State** | **Enrollment Institutions** | **Enrollment start date** | **Enrollment end date** |
| --- | --- | --- | --- |
| Arizona | Arizona State University,  Valleywise Health,  Phoenix Children’s Hospital | October 23, 2023 | April 30, 2024 |
| Michigan | University of Michigan,  Henry Ford Health | December 1, 2023 | April 25, 2024 |
| Missouri | Barnes Jewish Memorial Hospital,  St. Louis Children’s Hospital | October 10, 2023 | April 30, 2024 |
| Ohio | University Hospitals at Cleveland,  Cleveland Veterans’ Health Administration | November 27, 2023 | April 11, 2024 |
| Pennsylvania | University of Pittsburgh Medical Center,  Children’s Hospital of Pittsburgh | November 8, 2023 | April 25, 2024 |
| Texas | Baylor Scott & White Health | October 3, 2023 | April 30, 2024 |
| Washington | Kaiser Permanente Washington | October 11, 2023 | April 29, 2024 |

**Supplemental Figure 1.** Exclusion criteria figure
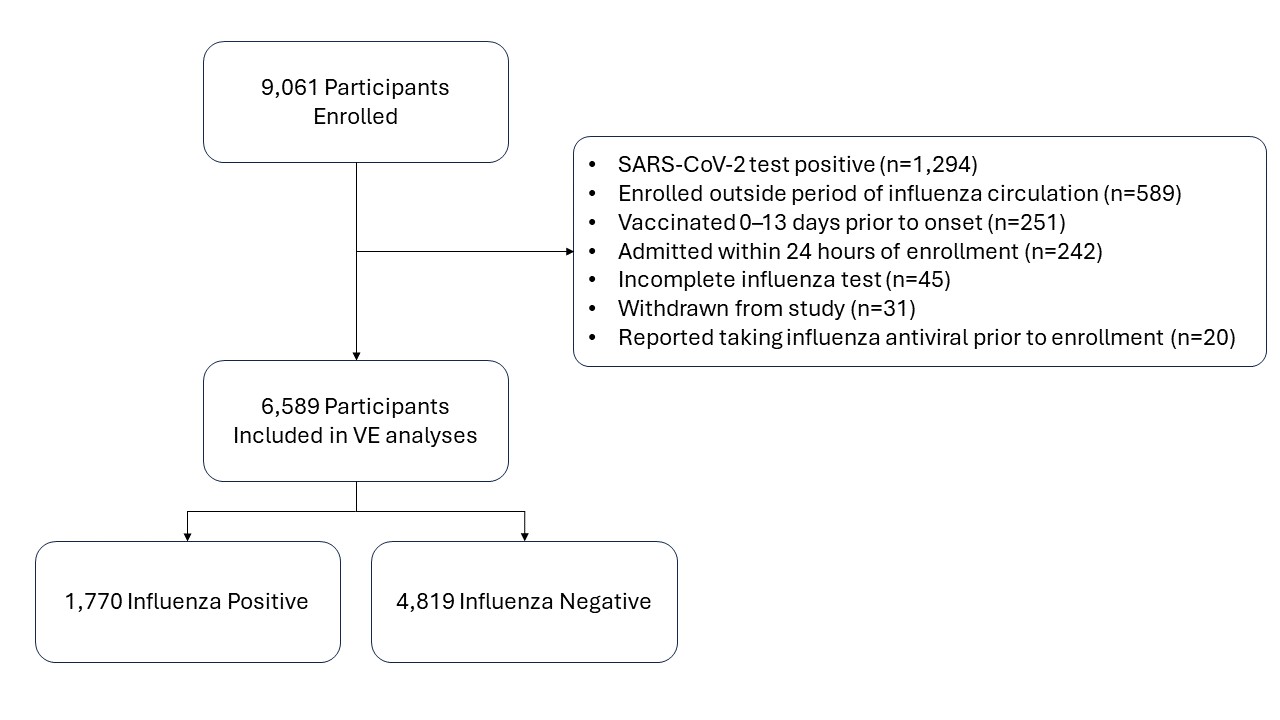

**Supplemental Figure 2.** Age group distribution of influenza A(H1N1)pdm09, A(H3N2), B/Victoria cases and influenza-negative control participants, October 2023 – April 2024.

|  | 8 mos - 8 years | 9-17 years | 18-49 years | 50-64 years | ≥65 years |
| --- | --- | --- | --- | --- | --- |
| A(H1N1)pdm09 (N=796) | 21 | 8 | 40 | 19 | 12 |
| A(H3N2) (N=323) | 13 | 8 | 60 | 11 | 8 |
| B/Victoria (N=563) | 31 | 22 | 42 | 4 | 1 |
| Negative (N=4819) | 19 | 8 | 40 | 16 | 16 |

**Supplemental Figure 3.** Influenza-positive cases by type/subtype and percent influenza positive by week of enrollment, October 2023 – April 2024.

Influenza virus detections in US Flu VE Network outpatients, 2023-24

N influenza-positive cases

Percent influenza positive

| Week of study enrollment | A(H1N1)pdm09 | A(H3N2) | B/Victoria | Percent influenza positive |
| --- | --- | --- | --- | --- |
| 2023 41 | 3 | 0 | 0 | 5.55556% |
| 2023 42 | 1 | 0 | 0 | 1.72414% |
| 2023 43 | 2 | 0 | 4 | 6.52174% |
| 2023 44 | 9 | 2 | 6 | 9.09091% |
| 2023 45 | 16 | 4 | 2 | 10.0457% |
| 2023 46 | 20 | 2 | 4 | 13.6126% |
| 2023 47 | 29 | 1 | 2 | 16.5829% |
| 2023 48 | 44 | 6 | 8 | 19.6141% |
| 2023 49 | 56 | 5 | 19 | 24.0469% |
| 2023 50 | 82 | 8 | 14 | 32.4159% |
| 2023 51 | 39 | 14 | 13 | 38.0435% |
| 2023 52 | 48 | 12 | 14 | 28.0576% |
| 2024 1 | 70 | 15 | 9 | 26.5928% |
| 2024 2 | 56 | 23 | 25 | 35.7143% |
| 2024 3 | 64 | 26 | 23 | 36.0759% |
| 2024 4 | 49 | 39 | 37 | 39.6923% |
| 2024 5 | 43 | 30 | 49 | 37.1601% |
| 2024 6 | 36 | 11 | 36 | 29.9652% |
| 2024 7 | 29 | 21 | 41 | 29.7468% |
| 2024 8 | 26 | 25 | 44 | 32.8904% |
| 2024 9 | 20 | 23 | 56 | 33.871% |
| 2024 10 | 17 | 12 | 37 | 29.4643% |
| 2024 11 | 7 | 11 | 24 | 19.1304% |
| 2024 12 | 8 | 12 | 12 | 17.1123% |
| 2024 13 | 5 | 7 | 22 | 20.8333% |
| 2024 14 | 5 | 5 | 12 | 14.9068% |
| 2024 15 | 0 | 5 | 5 | 8.0292% |
| 2024 16 | 6 | 2 | 4 | 11.4286% |
| 2024 17 | 2 | 1 | 2 | 7.8125% |
| 2024 18 | 1 | 0 | 0 |  |

**Supplemental Figure 4.** Sequenced A(H1N1)pdm09 virus genetic subclades by week of illness onset, October 2023 – April 2024

N of sequenced viruses

| Week of illness onset | 2a.1 | 2a |
| --- | --- | --- |
| 2023 41 | 2 | 0 |
| 2023 42 | 1 | 0 |
| 2023 43 | 2 | 0 |
| 2023 44 | 2 | 7 |
| 2023 45 | 10 | 5 |
| 2023 46 | 7 | 8 |
| 2023 47 | 14 | 11 |
| 2023 48 | 22 | 10 |
| 2023 49 | 40 | 10 |
| 2023 50 | 53 | 15 |
| 2023 51 | 24 | 11 |
| 2023 52 | 28 | 7 |
| 2024 1 | 41 | 13 |
| 2024 2 | 37 | 7 |
| 2024 3 | 43 | 11 |
| 2024 4 | 34 | 3 |
| 2024 5 | 29 | 2 |
| 2024 6 | 19 | 6 |
| 2024 7 | 21 | 2 |
| 2024 8 | 14 | 1 |
| 2024 9 | 9 | 2 |
| 2024 10 | 8 | 2 |
| 2024 11 | 2 | 1 |
| 2024 12 | 3 | 2 |
| 2024 13 | 1 | 0 |
| 2024 14 | 0 | 1 |
| 2024 16 | 3 | 0 |
| 2024 17 | 2 | 0 |

**Supplemental Table 2.** Adjusted vaccine effectiveness against any outpatient influenza-associated illness visits among patients aged ≥8 months enrolled at US Influenza Vaccine Effectiveness Network sites, October 2023 through April 2024 by time since vaccination.

| **Age group** | **Influenza Positive (Cases)** | | **Influenza Negative (Controls)** | | **Vaccine Effectiveness^a^** | |
| --- | --- | --- | --- | --- | --- | --- |
| Time since vaccination (days) | **Total** | **No. Vaccinated (%)** | **Total** | **No. Vaccinated (%)** | **VE %** | **(95% CI)** |
| **All ages ≥8 months** |  |  |  |  |  |  |
| 14–59 | 1486 | 58 (4) | 3566 | 491 (14) | 64 | (52 to 73) |
| 60–120 | 1587 | 159 (10) | 3771 | 696 (18) | 29 | (14 to 42) |
| >120 | 1551 | 123 (8) | 3634 | 559 (15) | 40 | (25 to 52) |
| **8 months –17 years** |  |  |  |  |  |  |
| 14–59 | 575 | 12 (2) | 1086 | 118 (11) | 83^b^ | (70 to 91) |
| 60–120 | 594 | 31 (5) | 1094 | 126 (12) | 51 | (27 to 69) |
| >120 | 582 | 19 (3) | 1070 | 102 (10) | 68^b^ | (48 to 81) |
| **18–64 years** |  |  |  |  |  |  |
| 14–59 | 839 | 36 (4) | 2097 | 257 (12) | 57 | (39 to 71) |
| 60–120 | 895 | 92 (10) | 2184 | 344 (16) | 23 | (0 to 40) |
| >120 | 873 | 70 (8) | 2111 | 271 (13) | 37 | (16 to 53) |
| **≥65 years** |  |  |  |  |  |  |
| 14–59 | 72 | 10 (14) | 383 | 116 (30) | 63^b^ | (28 to 83) |
| 60–120 | 98 | 36 (37) | 493 | 226 (46) | 15 | (-37 to 48) |
| >120 | 96 | 34 (35) | 453 | 186 (41) | 18 | (-35 to 51) |

Abbreviations: CI, confidence interval; VE, vaccine effectiveness

^a^ Models adjusted for study site, age, presence of ≥1 underlying health condition, and month of illness onset. 95% confidence intervals that exclude 0% are considered statistically significant

^b^ Unadjusted due to small sample size

**Supplemental Table 3**. Sensitivity analyses of vaccine effectiveness against any influenza among participants of all ages

|  | **Influenza Positive (Cases)** | | **Influenza Negative (Controls)** | | **Vaccine Effectiveness^a^** | |
| --- | --- | --- | --- | --- | --- | --- |
| **Sensitivity Analysis** | **Total** | **No. Vaccinated (%)** | **Total** | **No. Vaccinated (%)** | **VE %** | **(95% CI)** |
| **Primary analysis** | 1770 | 466 (26) | 4819 | 2023 (42) | 41 | (32 to 49) |
| **Varying exposure information source** |  |  |  |  |  |  |
| Defining vaccination status using self-reported information only | 1693 | 421 (25) | 4461 | 1710 (38) | 32 | (21 to 41) |
| Defining vaccination status using documented information only | 1801 | 340 (19) | 4927 | 1746 (35) | 44 | (35 to 52) |
| **Varying participant inclusion criteria** |  |  |  |  |  |  |
| Restricting to participants enrolled within 5 days of illness onset | 1605 | 407 (25) | 3937 | 1633 (41) | 41 | (32 to 49) |
| Including SARS-CoV-2-positive participants | 1803 | 473 (26) | 5825 | 2537 (44) | 45 | (37 to 52) |
| Removing participants from Arizona | 1564 | 454 (29) | 4448 | 1961 (44) | 40 | (30 to 47) |
| Removing participants from Michigan | 1688 | 432 (26) | 4534 | 1874 (41) | 42 | (33 to 49) |
| Removing participants from Missouri | 1425 | 379 (27) | 4385 | 1885 (43) | 45 | (37 to 53) |
| Removing participants from Ohio | 1513 | 405 (27) | 4310 | 1855 (43) | 43 | (34 to 51) |
| Removing participants from Pennsylvania | 1417 | 360 (25) | 3994 | 1617 (40) | 37 | (26 to 46) |
| Removing participants from Texas | 1419 | 367 (26) | 3372 | 1383 (41) | 38 | (28 to 47) |
| Removing participants from Washington | 1594 | 399 (25) | 3871 | 1563 (40) | 42 | (33 to 50) |

^a^ Models adjusted for study site, age, presence of ≥1 underlying health condition, and month of illness onset. 95% confidence intervals that exclude 0% are considered statistically significant
